# Determinants and Magnitude of Neonatal Sepsis at Hiwot Fana Comprehensive Specialized University Hospital, Harar, Ethiopia: A Cross-Sectional Study

**DOI:** 10.1101/2021.11.04.21265874

**Authors:** Astawus Alemayehu Feleke, Mohammed Yusuf Abdella, Abebaw Demissie W/Mariam

## Abstract

**Introduction:** Neonatal sepsis is a serious blood bacterial infection in neonates at the age of equal to or less than 28 days of life, and it’s still the major significant cause of death and long-term morbidity in developing countries. Therefore, this study has assessed the prevalence and related factors with neonatal sepsis among new born admitted to the neonatal intensive care unit at Hiwot Fana Comprehensive Specialized University Hospital, Harar, Ethiopia.

**Methods:** An institutional based retrospective cross-sectional study design was conducted among 386 neonates admitted to NICU from September 2017 to August 2019 G.C. A systematic random sampling method was used. Data was analyzed using SPSS V.26. Descriptive summary statistics was done. Bivariate analysis was computed to identify association between dependent and independent variables. Multivariate analysis was used to control possible confounder variables and variables with p-value <0.05 were declared as having statistically significant association.

**Result:** The prevalence of neonatal sepsis was 53.1% and 59.5% were males. Among the total neonates who had sepsis, 67.8% had early neonatal sepsis. Among neonatal factors, preterm neonates (AOR: 8.1, 95%CI: 2.1, 31.2), birth asphyxia (AOR: 4.7, 95%CI: 1.6, 13.6); and among maternal factors, urban residence (AOR: 0.26, 95%CI: 0.1, 0.5), ANC attendance (AOR: 0.32, 95%CI: 0.2, 0.6), SVD (AOR: 0.047, 95%CI: 0.01, 0.2), Maternal antibiotic use (AOR: 0.39; 95%CI: 0.2, 0.8), duration of rupture of membrane < 12 hours (AOR: 0.11; 95%CI: 0.05, 0.2) were found to have significant association with neonatal sepsis.

**Conclusion:** Overall, the magnitude of neonatal sepsis was high. Being preterm, low birth weight and having birth asphyxia were found to significantly increase the odds of neonatal sepsis. Urban residence, having ANC follow up, giving birth by SVD and CS, history of antibiotic use and having rupture of membrane < 18 hours were found to significantly decrease the odds of neonatal sepsis.

## Introduction

Sepsis is a systemic inflammatory reaction syndrome that occurs as a result of a suspected or confirmed infection. A dysregulated host response to infection causes a life-threatening organ failure. It’s also a prevalent last pathway to death from numerous infectious diseases around the world. ^1-3^ Neonatal sepsis is a serious blood bacterial infection in neonates at the age of equal to or less than 28 days of life, and it’s still the major significant cause of death and long-term morbidity in developing countries.^4, 5^

Globally, an estimated 1.3 to 3.9 million annual neonatal sepsis cases and 400 000 to 700 000 annual deaths worldwide. About 6,700 newborns die every day.^6^ Among hospital-born infants, hospital-acquired infections account for an estimated 4% to 56% of all deaths in the neonatal period. Among these, 84% of deaths are preventable.^5, 7^

Developing countries accounted for around 85.0 % of sepsis and deaths worldwide. Annually, an estimate of 5.29 to 8.73 million DALYs are lost due to neonatal sepsis in SSA countries.^8, 9^ The neonatal mortality rate in Ethiopia was 33 deaths per 1000 live births and the major causes of death in neonates are neonatal infections.^10-12^ Studies conducted in different parts of Ethiopia from 2015 – 2019 showed that the magnitude of neonatal sepsis ranged from 33.8-78% (33.8% in Wolayita, 64.8% in Gondar, 76.8% in Mekele, 77.8% in Shashamane, and 78.3% in Arba Minch). ^13-17^ The main significant factors associated with neonatal sepsis were maternal factors, (meconium stained amniotic fluid ^18, 19^, UTI during pregnancy ^12, 14, 15, 18, 19^, Place of delivery ^14, 15, 20^, Antenatal follow up ^19, 20^, and PROM ^12, 15, 17, 20, 21^) and neonatal factors (birth weight ^12, 17, 18^, and APGAR score ^17, 19, 21^).

As indicated by many studies conducted in different parts of Ethiopia, the magnitude of neonatal sepsis varies geographically. Thus, there is a need to perform periodic assessment of the problem in each location. In addition, in order to achieve the SDG goal of reducing neonatal mortality in Ethiopia, identifying factors contributing to neonatal sepsis is of paramount importance. There have been no studies in the research area that have investigated the magnitude and variables related with newborn sepsis to our knowledge. Thus, the prevalence of newborn sepsis and the factors that contribute to it have been investigated in this study among neonates admitted to the NICU at Hiwot Fana Comprehensive Specialized University Hospital.

## Method and Materials

### Study area and period

The study was conducted in Hiwot Fana Comprehensive Specialized University Hospital, which is located in Harar town, 526 km from Addis Ababa, the capital city of Ethiopia. Harar is one of the most popular historical towns. HFCSUH is a specialized university hospital established in 2006 and providing service to more than 5 million people in the catchment area. Currently, it is the only teaching and referral hospital in Harar city. Annually, around 1400 neonates are admitted to the NICU of HFCSUH.

### Study design

An institutional based retrospective cross-sectional study design was conducted.

### Source and study population

The source populations for the study were all neonates who were admitted and treated at the HFCSUH from September 2017 to August 2019 G.C. The study population was selected using systematic random sampling.

### Eligibility criteria

All neonates’ cards and age equal to or less than 28 days who were admitted to the NICU of HFCSUH were included and neonates’ cards with incomplete information were excluded.

### Sample size determination

A single population proportion formula was used to calculate the required sample size with the assumption of taking 95% confidence interval (CI), 5% marginal error and taking a proportion of 64.8 % from research done in Gondar, 2019.^14^

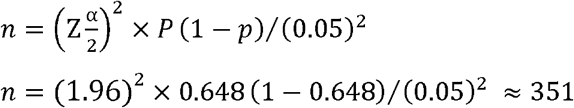

Where: n = required sample size; z = the standard normal deviation at 95% confidence interval=1.96; p = prevalence of neonatal sepsis 64.8% (0.648); d = margin of error 5% (0.05), by adding 10% (35.1) for non-response rate. Then, the total sample size to assess the prevalence of Neonatal Sepsis and associated factors was 386.

### Sampling technique and procedure

List of two year admitted neonates were taken from the database and K value was calculated as, K=N/n→ 2600/386=7 and sample were selected using systematic random sampling method by every k^th^ interval according to the registration order. Then the first number was selected by lottery method. So, every 7 intervals were selected until the completion of sample size. Incomplete sample was dropped out and replacement was done for it.

### Measurement

#### Study Variables

The dependent variable was neonatal sepsis and the independent variables included sociodemographic characteristics, maternal, and neonatal factors.

#### Data collection tools

After reviewing relevant literature, a semi structured data extraction tool was developed to obtain information on sociodemographic characteristics (age, sex, residence, and place of delivery); maternal factors (parity, mode of delivery, maternal diseases, maternal age, history of UTI, ANC follow-up, history of maternal fever, history of foul smelling liquor, history of chorioamniotis, meconium stained amniotic fluid, history of PROM, duration of rupture of membrane, duration of labor, and previous maternal use of medication); and Neonatal variables (birth weight, gestational age, birth asphyxia, and APGAR score).

#### Data processing and analysis

After the data was extracted from the card into the extraction sheet, it was checked manually for completeness and inconsistency. Then the data was coded and entered into Epidata v.3 and transported to SPSS version 24.0 for analysis. To describe the data, summary statistics were done. Bivariate regression analysis was used with a p-value <0.25 to check the existence of an association between each independent variable with the outcome variable. A multivariate logistic regression model was used to adjust the effects of possible cofounders on the outcome variable. Variables with a P-value < 0.05 were declared as having a statistically significant association.

#### Data Quality Assurance

First, a data extraction tool was prepared in English. Before starting the actual data collection, the quality of the data was assured by pre-testing the data extraction tool on a 10% of the total sample in Dilchora hospital, Dire Dawa. Training was given for data collectors for one day. On a daily basis, the collected data was reviewed by supervisors and authors to check for completeness and consistency before data entry.

### Operational definitions

#### Inborn

Neonate who was born in the maternity ward of HFCSUH and admitted to the NICU of HFCSUH.

#### Out born

Those neonates born outside of HFCSUH and admitted to the NICU of HFCSUH.

## Result

### Sociodemographic characteristics of neonate and mothers

A total of 386 neonate cards were reviewed, which is a 100% response rate. The majority of the mothers’ age ranged between 20 – 29 years. Regarding residence, 64.8% and 35.2% of mothers were from rural and urban, respectively. Out of 386, the majority, 234 (60.6%) of them were male. Regarding their age, the vast majority, 90.4%, were 0 – 7 days (Table 1).

**Table 1:** Sociodemographic characteristics of study participants in Hiwot Fana Comprehensive Specialized University Hospital, Harar, eastern Ethiopia, 2021

| Sociodemographic variables |  | Frequency (%) |
| --- | --- | --- |
| **Age of mother** | < 19 | 23 (6%) |
|  | 20 – 24 | 129 (33.4%) |
|  | 25 – 29 | 167 (43.3%) |
|  | 30 – 34 | 17 (4.4%) |
|  | ≥ 35 | 50 (13%) |
| **Residence** | Urban | 136 (35.2%) |
|  | Rural | 250 (64.8%) |
| **Age of neonate** | 0 – 7 days | 349 (90.4%) |
|  | 8 – 28 days | 37 (9.6%) |
| **Sex of neonate** | Male | 234 (60.6%) |
|  | Female | 152 (39.4%) |

### Maternal and neonatal health related characteristics

Out of 386 mothers of neonates, 228 (59.1%) of them had attended ANC during their pregnancy and 210 (54.4%) had a history of UTI. Of the total neonates, 233 (60.4%) of them were born in hospital. Most, 233 (60.4%) of them were born with SVD, and 270 (69%) of them were born at term. Regarding the duration of labour majority, 258 (66.3%) of neonates were born within less than 12 hours from the onset of labour. Among the total neonates, 235 (60.9%) were born with a weight of 2.5kg – 4kg. The initial assessment of neonates at birth indicated that the majority, 206 (53.4%) of them had < 7 APGAR score, while 242 (62.7%) and 104 (26.9%) of them had birth asphyxia and meconium-stained amniotic fluid, respectively (Table 2).

**Table 2:**
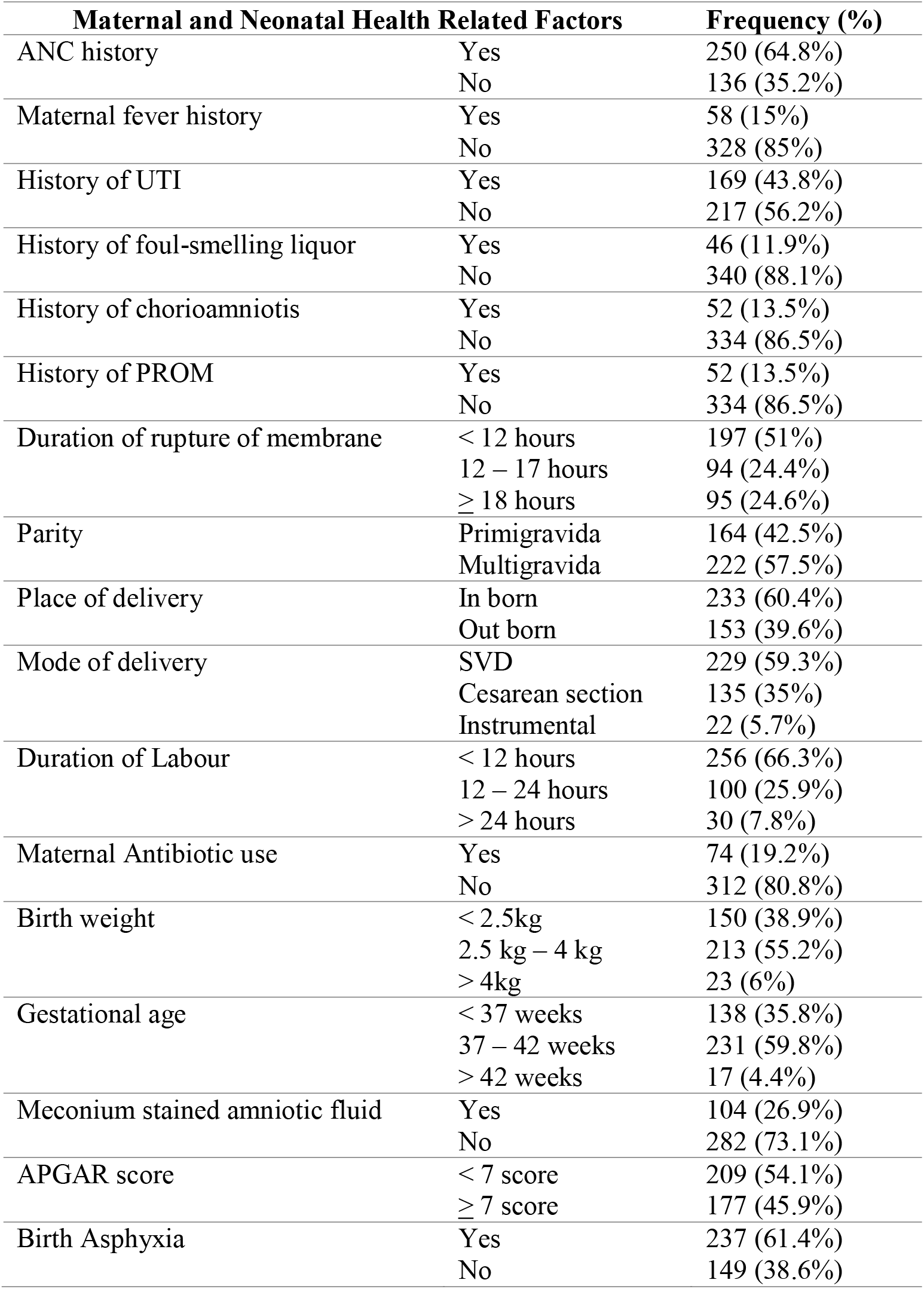
Maternal and neonatal health related factors among neonates admitted at Hiwot Fana Comprehensive Specialized University Hospital, Harar, eastern Ethiopia, 2021

#### Prevalence of Neonatal Sepsis Among Hospital Admitted Neonates

Out of the total 386 admitted neonates, 205 (53.1%) of them had neonatal sepsis (Figure 1). Among these, the majority, 139 (67.8%) of them had early onset neonatal sepsis and the rest had late onset of neonatal sepsis. Among the neonates who had sepsis, 122 (59.5%) were males, while 83 (40.5%) were females.

**Figure 1:**
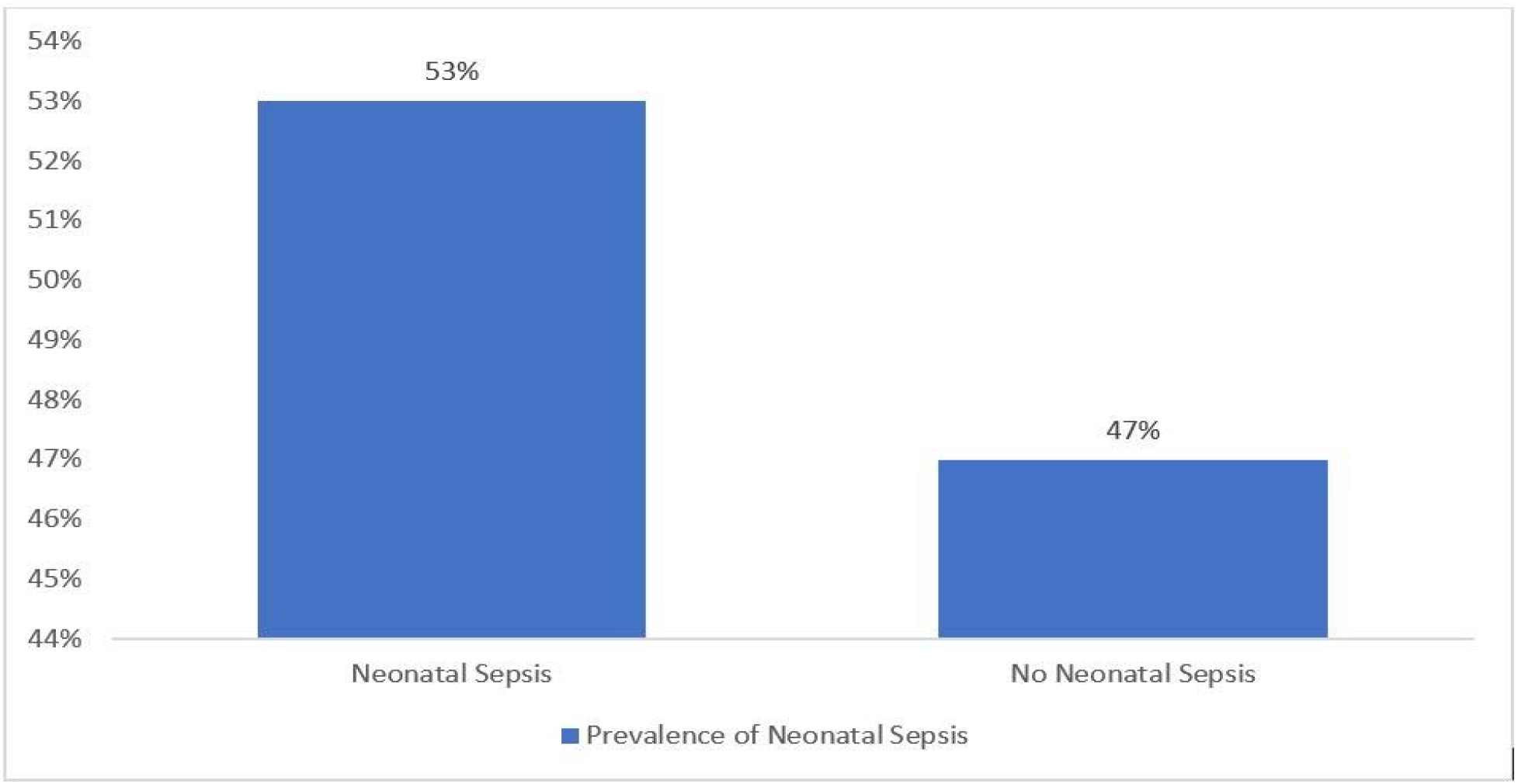
Prevalence of neonatal sepsis among neonates admitted in NICU at HFCSUH, Harar, Eastern Ethiopia, 2017-2019.

### Factors Associated with Neonatal Sepsis Among Hospital Admitted Neonates

In bivariate and multivariate analysis, neonatal and maternal factors were computed to assess factors associated with neonatal sepsis.

#### Neonatal related factors associated with neonatal sepsis

Preterm neonates were 8 (AOR: 8.1; 95% CI: 2.1,31.2) times more likely to develop neonatal sepsis than post term neonates. The odds of neonatal sepsis was 5 (AOR: 5.4; 95%CI: 1.5, 19.3) times among neonates born with < 2.5 kg than neonates born with > 4kg. Neonates with birth asphyxia were 4 (AOR: 4.7; 95%CI: 1.6, 13.6) times more likely to get sepsis than those who had no birth asphyxia (Table 3).

**Table 3:**
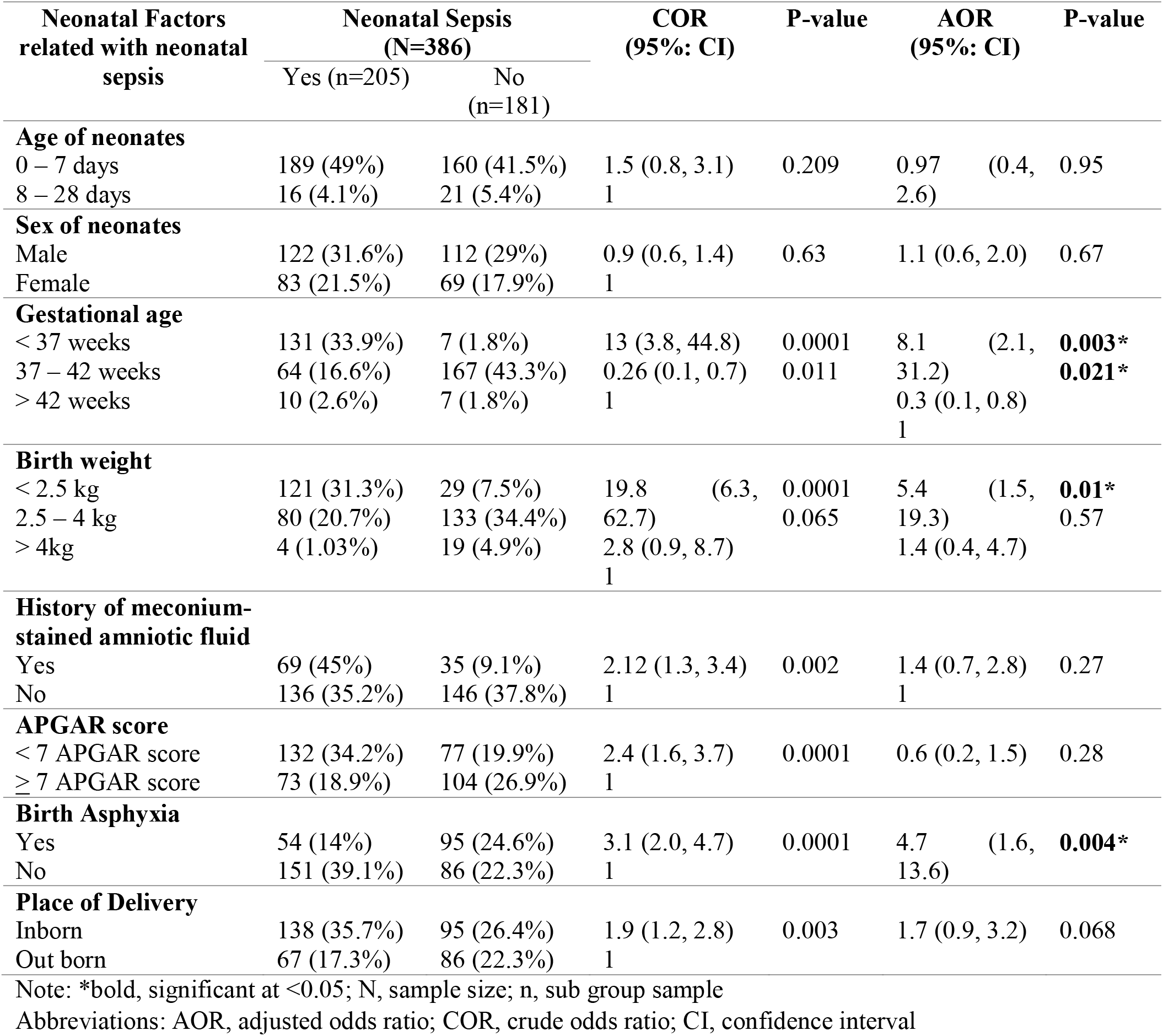
Factors associated with neonatal sepsis among neonates admitted in NICU at Hiwot Fana Comprehensive Specialized University Hospital, Harar, eastern Ethiopia, 2021

#### Maternal related factors associated with neonatal sepsis

Neonates born from mothers living in urban areas were 74% (AOR: 0.26; 95%CI: 0.1, 0.5) less likely to develop neonatal sepsis than neonates whose mothers lived in rural areas. Neonates born from mothers who had attended ANC were 68% (AOR: 0.32; 95%CI: 0.2, 0.6) less likely to develop neonatal sepsis than those neonates whose mothers had not attended ANC. Neonates born with spontaneous vaginal delivery were 95.3% (AOR: 0.047; 95%CI: 0.01, 0.2) less likely to develop neonatal sepsis than neonates born with instrumental delivery. Similarly, neonates born by cesarian section were 95% (AOR: 0.05; 95%CI: 0.01, 0.3) less likely to develop neonatal sepsis than neonates born by instrumental delivery.

Neonates born from mothers who had used antibiotics during pregnancy were 61% (AOR: 0.39; 95%CI: 0.2, 0.8) less likely to develop neonatal sepsis than neonates born from mothers who had not used antibiotics. Neonates born from mothers with a duration of rupture of membrane < 12 hours and 12 – 17 hours were 89% (AOR: 0.11; 95%CI: 0.05, 0.2), and 79% (AOR: 0.2; 95%CI: 0.1, 0.5) less likely to develop neonatal sepsis than neonates born from mothers with a duration of rupture of membrane > 18 hours, respectively (Table 4).

**Table 4:** Maternal factors associated with neonatal sepsis among neonates admitted in NICU at Hiwot Fana Comprehensive Specialized University Hospital, Harar, eastern Ethiopia, 2021

| Factor related with neonatal sepsis | Neonatal Sepsis (N=386) |  | COR (95%: CI) | P-value | AOR (95%: CI) | P-value |
| --- | --- | --- | --- | --- | --- | --- |
|  | Yes (n=205) | No (n=181) |  |  |  |  |
| **Age of mother** |  |  |  |  |  |  |
| < 19 | 12 (3.1%) | 11 (2.83%) | 1.1 (0.4, 2.9) | 0.86 | 1.44 (0.3, 6.3) | 0.625 |
| 20 – 24 | 66 (17.1%) | 63 (16.3%) | 1.05 (0.5, 2) | 0.88 | 1.45 (0.6, 3.8) | 0.449 |
| 25 – 29 | 92 (23.8%) | 75 (19.4%) | 1.2 (0.6, 2.3) | 0.52 | 1.9 (0.8, 4.4) | 0.142 |
| 30 – 34 | 10 (2.6%) | 7 (1.8%) | 1.4 (0.5, 4.3) | 0.53 | 1.2 (0.3, 5.1) | 0.83 |
| ≥ 35 | 25 (6.5%) | 25 (6.5%) | 1 |  | 1 |  |
| **Residence** |  |  |  |  |  |  |
| Urban | 48 (12.4%) | 88 (22.8%) | 0.3 (0.2, 0.5) | 0.0001 | 0.26 (0.1, 0.5) | **0.0001\*** |
| Rural | 157 (40.7%) | 93 (24.1%) | 1 |  | 1 |  |
| **History of ANC** |  |  |  |  |  |  |
| Yes | 112 (29%) | 138 (35.8%) | 0.3 (0.2, 0.6) | 0.0001 | 0.32 (0.2, 0.6) | **0.0001\*** |
| No | 93 (24.1%) | 43 (11.1%) | 1 |  | 1 |  |
| **History of maternal fever** |  |  |  |  |  |  |
| Yes | 40 (10.4%) | 18 (4.7%) | 1 |  | 1 |  |
| No | 165 (42.7%) | 163 (42.2%) | 0.46 (0.2, 0.8) | 0.01 | 1.3 (0.5, 3.4) | 0.546 |
| **History of UTI** |  |  |  |  |  |  |
| Yes | 116 (30.1%) | 53 (13.7%) | 3.1 (2.1, 4.8) | 0.0001 | 3.6 (2.1, 6.1) | **0.0001\*** |
| No | 89 (23%) | 128 (33.2%) | 1 |  | 1 |  |
| **History of foul smelling** |  |  |  |  |  |  |
| Yes | 33 (8.5%) | 13 (3.4%) | 1 |  | 1 |  |
| No | 172 (44.6%) | 168 (43.5%) | 0.4 (0.2, 0.8) | 0.008 | 0.4 (0.1, 1.3) | 0.125 |
| **History of chorioamniotitis** |  |  |  |  |  |  |
| Yes | 40 (10.4%) | 12 (3.1%) | 1 |  | 1 |  |
| No | 165 (42.7%) | 169 (43.8%) | 0.3 (0.1, 0.6) | 0.0001 | 0.34 (0.1, 1.1) | 0.079 |
| **History of PROM** |  |  |  |  |  |  |
| Yes | 29 (7.5%) | 23 (6%) | 1 |  | 1 |  |
| No | 176 (45.6%) | 158 (40.9%) | 0.9 (0.5, 1.6) | 0.68 | 1.7 (0.7, 4.1) | 0.254 |
| **Duration of labour** |  |  |  |  |  |  |
| < 12 hours | 134 (34.8%) | 122 (31.6%) | 1.2 (0.6, 2.6) | 0.55 | 1.9 (0.6, 5.3) | 0.24 |
| 12 – 24 hours | 57 (14.8%) | 43 (11.1%) | 1.5 (0.6, 3.4) | 0.32 | 1.6 (0.5, 4.7) | 0.433 |
| > 24 hours | 14 (3.6%) | 16 (4.3%) | 1 |  | 1 |  |
| **Parity** |  |  |  |  |  |  |
| Primigravida | 87 (22.5%) | 77 (20%) | 0.9 (0.7, 1.5) | 0.98 | 1.1 (0.6, 2.1) | 0.79 |
| Multigravida | 118 (30.6%) | 104 (26.9%) | 1 |  | 1 |  |
| **Mode of delivery** |  |  |  |  |  |  |
| SVD | 122 (31.6%) | 107 (27.7%) | 0.11 (0.03, 0.5) | 0.004 | 0.047(0.01, 0.2) | **0.0001\*** |
| Cesarean section | 63 (16.3%) | 72 (18.7%) | 0.09 (0.02, 0.4) | 0.001 | 0.05 (0.01, 0.3) | **0.001\*** |
| Instrumental delivery | 20 (5.2 %) | 2 (0.52%) | 1 |  | 1 |  |
| **Maternal antibiotic use** |  |  |  |  |  |  |
| Yes | 27 (7%) | 47 (12.2%) | 0.43 (0.3, 0.7) | 0.002 | 0.39 (0.2, 0.8) | **0.0001\*** |
| No | 178 (46.1%) | 134 (34.7%) | 1 |  | 1 |  |
| **Duration of rapture of membrane** |  |  |  |  |  |  |
| < 12 hours | 76 (19.7%) | 121 (31.3%) | 0.11 (0.6, 0.2) | 0.0001 | 0.11 (0.05, 0.2) | **0.0001\*** |
| 12 – 17 hours | 48 (12.4%) | 46 (11.9%) | 0.18 (0.1, 0.4) | 0.0001 |  | **0.0001\*** |
| ≥ 18 hours | 81 (21%) | 14 (3.6%) | 1 |  | 0.21 (0.1, 0.5) |  |
|  |  |  |  |  | 1 |  |

Note: \*bold, significant at <0.05; N, sample size; n, sub group sample
Abbreviations: AOR, adjusted odds ratio; COR, crude odds ratio; CI, confidence interval

## Discussion

In our findings, the prevalence of neonatal sepsis was 53.1%. Other similar studies conducted in different parts of Ethiopia indicated a higher prevalence of neonatal sepsis, 78.3% in Arba Minch.^17^, 77.9% in Shashamane^16^, 76.8% in Mekelle^15^, 64.8% in Gondar.^14^ The reason for the low prevalence of neonatal sepsis in our finding compared to the above studies could be due to difference in population variation and due to the fact that our study was conducted in a specialized university referral hospital which has relatively better facilities and human resources. On the contrary, our study finding was higher than the finding reported from Wolaita Sodo, 33.8%^13^, These differences might be due to sociodemographic variation and sample size differences.

In our study, the prevalence of neonatal sepsis was higher among males than females 31.6%. This finding was consistent with studies conducted in Bahar Dar, Gondar and Arba Minch, 64%^22^, 41.1%^14^, and 58.2%^17^, respectively.

The prevalence of neonatal sepsis in this study was found to be higher, 49% among early neonates (0-7 days). A similar finding was also reported from other studies in West Showa, Arba Minch, Bahar Dar, Gondar, and Wolaita Sodo, which reported 94.9%^20^, 84%^17^, 72.9%^22^, 48.2%^14^, and 40.4%^13^, respectively.

In our finding, preterm neonates were 8 times more likely to develop neonatal sepsis than post term neonates. This is consistent with study conducted in Debre Markos where preterm neonates were found to be 6 times more likely to develop neonatal sepsis. In addition, other studies also reported similar findings.^21, 23, 24^

Our study revealed that the odds of neonatal sepsis was 5 times higher among neonates born with low birth weight. This is consistent with studies conducted in different parts of Ethiopia, in Durame, Arsi, Arba Minch, Jimma, and Gondar. ^12, 17, 18, 24, 25^

According to our study, neonates with birth asphyxia was 4 times more likely to develop neonatal sepsis than those who had no birth asphyxia. This is consistent with a study conducted in Shashamane.^16^

In our study, neonates born from mothers who had ANC follow up were 68% less likely to develop neonatal sepsis than those neonates whose mothers had not ANC follow up. This result was consistent with study conducted in west Shewa, Oromia Region.^20^

Our study revealed that, neonates born from mothers who had a history of UTI were 3 times more likely to develop neonatal sepsis than neonates born from mothers who had no history of UTI. This finding was consistent with similar studies conducted in Ethiopia; in Durame^18^, Gondar^14^ and Mekelle.^15^

Our study findings indicated that neonates who were born by SVD and Cesarian section were found to be 95.3% and 95% less likely to get neonatal sepsis than children born by instrumental delivery, respectively. This finding was contradictory to the finding reported in Gondar, which reported that neonates born by cesarian section were more likely to develop neonatal sepsis than neonates born by SVD and instrumental delivery.^25^ This difference could be due to a difference in population and sample size variation.

In our study, neonates born of mothers who had a duration of rupture membrane < 12 hours and 12 – 17 hours were 89% and 79% less likely to have neonatal sepsis than those neonates born of mothers with a duration of rupture of membrane ≥ 18 hours. Similar findings were also reported from studies in Arba Minch^17^ and Debre Markos, Ethiopia^21^.

In our study, neonates born from mothers who had a history of antibiotic use were 61% less likely to have neonatal sepsis than those born from mothers who had no history of antibiotic use.

Similarly, neonates born from mothers who lived in urban areas were 74% less likely to have neonatal sepsis than neonates born from mothers living in rural areas. These two factors were the only variables which had a significant association with neonatal sepsis and which were not reported in other similar studies.

## Conclusion

Overall, the magnitude of neonatal sepsis was high and it was more prevalent among male neonates. Among neonatal factors; being preterm, low birth weight and having birth asphyxia were found to significantly increase the odds of neonatal sepsis. Among maternal factors; urban residence, having ANC follow up, give birth by SVD and CS, history of antibiotic use and having rupture of membrane < 18 hours were found to significantly decrease the odds of neonatal sepsis.

Provision of neonatal and obstetrics care as per standard during in pre, intra and postnatal period. Training of health professionals on infection prevention and safe delivery practice should be provided.

## Limitation of the study

The research was conducted using secondary data, which is difficult to generalize to the general population. The study used a cross-sectional study design. Therefore, there is a temporal issue.

## Data Availability

All data produced in the present study are available upon reasonable request to the authors

## Abbreviations

CEO: Chief Executive Officer
CS: Cesarian Section
NICU: Neonatal Intensive Care Unit
HFCSUH: Hiwot Fana Comprehensive Specialized University Hospital
IHREC: Institutional Health Research Ethical Committee
PROM: Premature Rupture of Membrane
SDGs: Sustainable Developmental Goals
SVD: Spontaneous Vaginal Delivery
SPSSs: Statistical Package for Social Science
SSA: sub-Saharan Africa

## Acknowledgments

We thank Almighty God. Also, we would like to extend thanks to Hiwot Fana Comprehensive specialized University Hospital.

## Disclosure

### Ethics approval

Ethical Approval letter was obtained from Rift Valley University IHREC with reference no. IHREC 0015/02/01/2020, and consent was obtained to collect data. Confidentiality of all information had been maintained.

### Consent to participate

Not applicable

### Consent for publication

Not applicable

### Data availability

Any time, the corresponding author provides an additional resource on request.

### Author contributions

AA MY, and AD participated in the study from inception to design, acquisition of data, analysis, and interpretation of the results.

## Reference

1. Geme, R.M.K.J.S., Nelson Textbook of Pediatrics, 2-Volume Set. 2019: Publisher: Elsevier.

2. Singer, M., et al., The third international consensus definitions for sepsis and septic shock (Sepsis-3). Jama, 2016. 315(8): p. 801–810.

3. WHO. Sepsis. WHO Sepsis Response 2020 [cited 2020; the Seventieth World Health Assembly adopted Resolution]. Available from: https://www.who.int/news-room/fact-sheets/detail/sepsis

4. Seale, A.C., et al., Estimates of possible severe bacterial infection in neonates in sub-Saharan Africa, south Asia, and Latin America for 2012: a systematic review and meta-analysis. Lancet Infect Dis, 2014. 14(8): p. 731–741.

5. Organization, W.H., Global report on the epidemiology and burden of sepsis: Current evidence, identifying gaps and future directions. 2020.

6. IGME), U.N.I.-a.G.f.C.M.E.U., Levels and Trends in Child Mortality: Report 2019, in estimates developed by the United Nations Inter-agency Group for Child Mortality Estimation. 2020: New York, USA.

7. Lawn, J.E., et al., Every Newborn: progress, priorities, and potential beyond survival. Lancet, 2014. 384(9938): p. 189–205.

8. Rudd, K.E., et al., Global, regional, and national sepsis incidence and mortality, 1990–2017: analysis for the Global Burden of Disease Study. The Lancet, 2020. 395(10219): p. 200–211.

9. Ranjeva, S.L., B.C. Warf, and S.J. Schiff, Economic burden of neonatal sepsis in sub-Saharan Africa. BMJ Glob Health, 2018. 3(1): p. e000347.

10. Ethiopian Public Health Institute, E., F. Federal Ministry of Health, and Icf, Ethiopia Mini Demographic and Health Survey 2019. 2021, EPHI/FMoH/ICF: Addis Ababa, Ethiopia.

11. Central Statistical Agency (CSA), a.I., Ethiopia Demographic and Health Survey 2016: Key Indicators Report. 2016, CSA and ICF: Addis Ababa, Ethiopia, and Rockville, Maryland, USA.

12. Debelew, G.T., M.F. Afework, and A.W. Yalew, Determinants and causes of neonatal mortality in Jimma zone, southwest Ethiopia: a multilevel analysis of prospective follow up study. PloS one, 2014. 9(9): p. e107184.

13. Mersha, A., et al., Neonatal sepsis and associated factors among newborns in hospitals of Wolaita Sodo Town, Southern Ethiopia. Research and reports in neonatology, 2019. 9: p. 1.

14. Agnche, Z., H.Y. Yeshita, and K.A. Gonete, Neonatal sepsis and its associated factors among neonates admitted to neonatal intensive care units in primary hospitals in central gondar zone, northwest ethiopia, 2019. Infection and Drug Resistance, 2020. 13: p. 3957.

15. Gebremedhin, D., H. Berhe, and K. Gebrekirstos, Risk factors for neonatal sepsis in public hospitals of Mekelle City, North Ethiopia, 2015: unmatched case control study. PloS one, 2016. 11(5): p. e0154798.

16. Getabelew, A., et al., Prevalence of neonatal sepsis and associated factors among neonates in neonatal intensive care unit at selected governmental hospitals in Shashemene Town, Oromia Regional State, Ethiopia, 2017. International journal of pediatrics, 2018. 2018.

17. Mustefa, A., A. Abera, and A. Aseffa, Prevalence of neonatal sepsis and associated factors amongst neonates admitted in arbaminch general hospital, arbaminch, southern Ethiopia, 2019. J Pediatr Neonatal Care, 2020. 10(1): p. 1–7.

18. Dirirsa, D.E., B. Dibaba Degefa, and A.D. Gonfa, Determinants of neonatal sepsis among neonates delivered in Southwest Ethiopia 2018: A case-control study. SAGE Open Medicine, 2021. 9: p. 20503121211027044.

19. Akalu, T.Y., et al., Predictors of neonatal sepsis in public referral hospitals, Northwest Ethiopia: A case control study. Plos one, 2020. 15(6): p. e0234472.

20. Bulto, G.A., et al., Determinants of Neonatal Sepsis among Neonates Admitted to Public Hospitals in Central Ethiopia: Unmatched Case-control Study. Global Pediatric Health, 2021. 8: p. 2333794X211026186.

21. Alemu, M., et al., Determinants of neonatal sepsis among neonates in the northwest part of Ethiopia: case-control study. Italian journal of pediatrics, 2019. 45(1): p. 1–8.

22. Tewabe, T., et al., Clinical outcome and risk factors of neonatal sepsis among neonates in Felege Hiwot referral Hospital, Bahir Dar, Amhara Regional State, North West Ethiopia 2016: a retrospective chart review. BMC Research Notes, 2017. 10(1): p. 265.

23. Gebrehiwot, A., et al., Bacterial profile and drug susceptibility pattern of neonatal sepsis in Gondar University Hospital, Gondar Northwest Ethiopia. Der Pharmacia Lettre, 2012. 4(6): p. 1811–1816.

24. Sorsa, A., Epidemiology of neonatal sepsis and associated factors implicated: observational study at neonatal intensive care unit of Arsi University Teaching and Referral Hospital, South East Ethiopia. Ethiopian journal of health sciences, 2019. 29(3).

25. Moges, F., et al., Bacterial etiologic agents causing neonatal sepsis and associated risk factors in Gondar, Northwest Ethiopia. BMC pediatrics, 2017. 17(1): p. 1–10.

